# Respiratory Viral Sequencing Panel identifies SARS-CoV-2 variants, transmission and other co-circulating viruses in Georgia, USA: A Diagnostic and Epidemiologic Tool for Mass Surveillance in COVID-19 Pandemic

**DOI:** 10.1101/2021.07.24.21261046

**Authors:** Nikhil S Sahajpal, Ashis K Mondal, Allan Njau, Zachary Petty, Jiani Chen, Sudha Ananth, Pankaj Ahluwalia, Colin Williams, Ted M Ross, Alka Chaubey, Grace DeSantis, Gary P. Schroth, Justin Bahl, Ravindra Kolhe

**Affiliations:** Department of Pathology, Medical College of Georgia, Augusta University, GA, U.S.A.; Department of Pathology, Aga Khan University Hospital, Nairobi, Kenya; Center for Ecology of Infectious Diseases, Institute of Bioinformatics, University of Georgia, Athens, GA, USA; Center for Vaccines and Immunology, University of Georgia, GA, U.S.A.; Bioano Genomics Inc., San Diego, CA, U.S.A; Research and Development, Illumina Inc, San Diego, CA, U.S.A.; Department of Infectious Disease, University of Georgia, Athens, GA, USA; Department of Epidemiology and Biostatistics, University of Georgia, Athens, GA, USA

## Abstract

**Background:** In the current phase of COVID-19 pandemic, we are facing two serious public health challenges that include deficits in SARS-CoV-2 variant monitoring, and neglect of other co-circulating respiratory viruses. Additionally, accurate assessment of the evolution, extent and dynamics of the outbreak are required to understand the transmission of the virus amongst seemingly unrelated cases and provide critical epidemiological information. To address these challenges, we evaluated a new high-throughput next-generation sequencing (NGS), respiratory viral panel (RVP) that includes 40 viral pathogens with the aim of analyzing viral subtypes, mutational variants of SARS-CoV-2, model to understand the spread of the virus in the state of Georgia, USA, and to assess other circulating viruses in the same population.

**Methods:** This study evaluated a total of 522 samples that included 483 patient samples and 42 synthetic positive control material. The performance metrics were calculated for both clinical and reference control samples by comparing detection results with the RT-PCR assay. The limit of detection (LoD) studies were conducted as per the FDA guidelines. Inference and visualization of the phylogeny of the SARS-CoV-2 sequences were performed through the Nextstrain Command-Line Interface (CLI) tool, utilizing the associated augur and auspice toolkits.

**Results:** The performance metrics calculated using both the clinical samples and the reference controls revealed a PPA, NPA and accuracy of 95.98%, 85.96% and 94.4%, respectively. The LoD was determined to be 10 copies/ml with all 25 replicates detected across two different runs. The clade for pangolin lineage B that contains certain distant variants, including P4715L in ORF1ab, Q57H in ORF 3a and, S84L in ORF8 covarying with the D614G spike protein mutation were the most prevalent, early in the pandemic, in Georgia, USA. In our analysis, isolates from the same county formed paraphyletic groups, which indicated virus transmission between counties.

**Conclusion:** The study demonstrates the clinical and public health utility of the NGS-RVP to identify novel variants that can provide actionable information to prevent or mitigate emerging viral threats, models that provide insights into viral transmission patterns and predict transmission/ resurgence of regional outbreaks and provide critical information on co-circulating respiratory viruses that might be independent factors contributing to the global disease burden.

## Introduction

The global society reeled back by the COVID- 19 pandemic, now in its second year, has seen over 150 M cases and over 3.1 M COVID-19 related deaths. (https://coronavirus.jhu.edu/map.html, last accessed 05/02/2021). Conversely, united efforts around the world in cultural economic, and scientific realms; notably high throughput diagnostic solutions, therapeutic options, and more recently, the massive rollout of vaccination, with more than 450M people have received at least one dose (https://coronavirus.jhu.edu/map.html, last accessed 05/02/2021), is showing promise towards control of the pandemic. With many countries, having weathered several waves of the pandemic, and with spikes still being anticipated in the future, persistent effort and surveillance are still required. Human activity and variation in viral subtypes that determine transmissibility and pathogenicity have largely been implicated for these trends. Several SARS-CoV-2 variants have been identified, but three recently reported strains are of significant concern. The 20B/501Y.V1 or VOC 202012/01 variant of the B.1.1.7 (alpha) lineage that is defined by 17 mutations (14 non-synonymous mutations and 3 deletions) was identified in the UK [**1**,**2**], the 20C/501Y.V2 strain that emerged independently of the B.1.17 to B1.351 (beta) lineage was identified in South Africa, and B.1.617.2 (delta) variant has emerged most recently [**3**,**4**]. The VOC 202012/01 SARS-CoV-2 strain was 43-90% more transmissible compared to the preexisting strains and led to “Tier 4” restrictions in December 2020 in the UK, and the delta variant is emerging as a similar threat [**5**]. The emergence of these novel strains imposes the need to sequence the SARS-CoV-2 genome in clinical laboratories across the entire globe as any lacunae in monitoring the variation in the SARS-CoV-2 genome can lead to serious public health consequences. In addition, another immediate public health deficit pertaining to other respiratory viruses is already of major concern, since co-circulating respiratory viruses other than SARS-CoV-2 have largely remained undocumented for nearly the entire year of 2020.

In the past, pandemics due to novel pathogens have amplified the incidence of respiratory tract infections (RTI) leading to morbidity and mortality exceeding the seasonal levels of the disease. The Global Burden of Disease (2017) data demonstrated that influenza contributed 11.5 % of the total lower respiratory tract infections (LRTIs), leading to over 9 million hospitalizations and 145,000 deaths across all age groups [**6**]. Other viral pathogens including Rhinoviruses, Parainfluenza viruses, Respiratory Syncytial virus, and Adenoviruses also account for respiratory tract infections of varying severity, as either mono-infections or coinfections. Coinfection with viral, bacterial, or fungal pathogens has been associated with disease severity and death in the current pandemic [**7**]. A meta-analysis including 30 studies and 3834 COVID-19 patients of all age groups and settings found that 7% of hospitalized patients had bacterial co-infection, with the most common being *Mycoplasma pneumonia, Pseudomonas aeruginosa*, and *Haemophilus influenza*; and viral co-infections were identified in 3% of the patients [**8**]. Another meta-analysis found a slightly higher viral coinfection rate of 7% [**9**]. A study conducted in California found a co-infection rate of 20.7% among SARS-CoV-2 positive patients [**10**], while this was 3.3% in a Chicago study [**11**]. In both of these studies, Rhinovirus and enterovirus were the commonest. In addition to multiplex RT-PCR assays used to identify co-infections, metagenomics sequencing has demonstrated utility in unbiased identification of co-infection or colonization [**12, 13**].

Owing to the diversion of resources and supplies to SARS-CoV-2 testing, testing of viral pathogens that normally cause seasonal RTI has been largely neglected. The current practice has posed serious public health gaps both at a clinical and epidemiological level, especially now that the transmission and infective mutations are emerging, and the virus is persisting to more transmissible variants. Further, as the COVID-19 vaccination process is underway, the screening of COVID-19 by RT-PCR-based SARS-CoV-2 detection methods needs to be complemented with two additional monitoring measures. The first is sequencing the SARS-CoV-2 genome to identify novel variants that can provide actionable information to prevent or mitigate emerging viral threats and the second is to test for co-circulating respiratory viruses that might be independent factors contributing to the global disease burden. The use of multiple antimicrobial agents in moderate-severely ill COVID-19 patients can also be rationalized based on such studies. To address these clinical and public health challenges, we aimed to evaluate the performance of a new high throughput next-generation sequencing respirator viral panel (NGS-RVP) that includes 40 viral pathogens: and analyze viral subtypes; mutational variants of SARS-CoV-2 in the state of Georgia, USA; develop models to understand the spread of the virus in the state of Georgia, and to assess the other circulating viruses in the same population.

## Materials and Methods

### Study site and ethics

The study was performed at Augusta University, GA, USA under IRB approval. This site is CLIA accredited laboratory for high complexity testing and is one of the main SAR-CoV-2 testing centers in the state. The samples were processed under an approved HAC by the IRB Committee A (IRB registration # 611298), Augusta University, GA. Based on the IRB approval, the need for consent was waived, all PHI was removed and the data was anonymized before accessing for the study.

### Samples

This study evaluated a total of 522 samples that included 483 patient samples, 39 synthetic positive control material (Twist biosciences), and 3 no-template controls (NTC). The 483 patient samples were previously tested for SAR-CoV-2 by RT-PCR-based COVID-19 diagnostic assay (PerkinElmer Inc. assay (LoD 20 copies/ml). Of the 483 samples, 471 were NPS and 12 were saliva samples with 398 positive and 85 negatives for SAR-CoV-2 by RT-PCR. For temporal distribution, selected samples included those collected from the month of March through October 2020. In addition, to ensure samples from the entire state were represented, the state of Georgia was divided into three arbitrary regions including north, southwest, and southeast Georgia, with 31, 72, and 380 samples selected from each of the regions, respectively. Relevant meta-data including age sex and ethnicity were recorded. Positive samples with a wide range of Ct values (*N*: 6.9–36.8, *ORF1ab*: 8.7-39.6) were chosen. Thirty-nine (39) sample dilutions of synthetic positive control material with 10^7^ copies/ml, 10^6^ copies/ml, 90 copies/ml, 30 copies/ml and 10 copies/ml were included.

### Laboratory processes

#### RNA extraction and RT-PCR for SARS-CoV-2

All patient samples were tested for SARS-CoV-2 using an assay based on RNA extraction followed by TaqMan-based RT-PCR assay to conduct in vitro transcription of SARS-CoV-2 RNA, DNA amplification, and fluorescence detection (PerkinElmer Inc, USA). The assay targets specific genomic regions of SARS-CoV-2: nucleocapsid (*N*) gene and *ORF1ab* with an RNA internal control (IC, bacteriophage MS2) to monitor the processes from nucleic acid extraction to fluorescence detection. The probes are labeled with FAM, ROX, and VIC dyes, respectively, to differentiate the fluorescent signals from each target. The assay validation was performed as per FDA guidelines, following the manufacturer’s protocol. In brief, 300µl sample was used for RNA extraction (chemagic 360 instrument, PerkinElmer Inc), to which 5µl internal control, 4µl Poly(A) RNA, 10µl proteinase K, and 300µl lysis buffer 1 were added. From 60µl eluate, the RT-PCR reaction was set up, which included 10µl of extracted nucleic acid and 5µl of PCR master mix. PCR was performed using QuantStudio 3 and 5 Real-Time PCR Systems (Thermo Fisher Scientific). The LoD of this assay is 20 copies/ml.

#### Next-generation sequencing

Library preparation was performed following the Illumina RNA Prep with Enrichment kits which leverage BLT technology paired with fast enrichment (cat number 20040537, Illumina). Briefly, 8.5 uL of extracted RNA, by the methodology described above, was denatured followed by first and second-strand DNA synthesis. This was followed by tagmentation, which uses enrichment bead-linked transposomes (BLT) to tagment double-stranded cDNA. This process fragments cDNA and adds adapter sequences. After tagmentation, the fragments were purified and amplified to add index adapter sequences for dual indexing, and P7 and P5 sequences for clustering. Four index sets, A, B, C, and D each containing 96 unique, single-use Illumina® DNA/RNA UD Indexes were used. Following clean-up, libraries were quantified using Invitrogen Qubit dsDNA broad range Assay Kit (Thermo Fischer Scientific). Subsequently, 7.5ul of the library was used for hybridization using oligos from the respiratory viral panel. This was followed by bead-based capture of hybridized probes, amplification, clean-up, and quantification of the enriched library. The viruses targeted by the Respiratory Virus Oligos Panel V2 (RVOP v2, cat number 20044311, Illumina) are shown in **Table 1**. As an additional QC check, representative libraries were analyzed for fragment size using QIaxel (QIAGEN, Germany). Normalized libraries diluted to an equimolar concentration of 0.8pM were then pooled into three runs. Using a 150bp paired-end sequencing approach and 300 cycles, the libraries were sequenced on the NextSeq 500/550 high-throughput sequencer using a V2 flow cell kit (Illumina).

**Table 1.** Table showing the viruses included in the enrichment workflow of the Respiratory Virus Oligos Panel V2.

| S. No. | Virus |
| --- | --- |
| 1 | Human Corona virus 229E |
| 2 | Human Corona virus NL63 |
| 3 | Human Corona virus OC43 |
| 4 | Human Corona virus HKU1 |
| 5 | SARS-CoV-2 |
| 6 | Human adenovirus B1 |
| 7 | Human adenovirus C2 |
| 8 | Human adenovirus E4 |
| 9 | Human bocavirus 1 (Primate bocaparvovirus 1 isolate st2) |
| 10 | Human bocavirus 2c PK isolate PK-5510 |
| 11 | Human bocavirus 3 |
| 12 | Human parainfluenza virus 1 |
| 13 | Human parainfluenza virus 2 |
| 14 | Human parainfluenza virus 3 |
| 15 | Human parainfluenza virus 4a |
| 16 | Human metapneumovirus (CAN97-83) |
| 17 | Respiratory syncytial virus (type A) |
| 18 | Respiratory syncytial virus 9320 (type B) |
| 19 | Influenza A virus (A/Puerto Rico/B/1934(H1N1)) |
| 20 | Influenza A virus (A/Korea/426/1968(H2N2)) |
| 21 | Influenza A virus (A/New York/392/2004(H3N2)) |
| 22 | Influenza A virus (A/goose/Guangdong/1/1996(H5N1)) |
| 23 | Human bocavirus 4 NI strain HBoV4-NI-385 |
| 24 | KI Polyomavirus Stockholm 60 |
| 25 | WU Polyomavirus |
| 26 | Human Parechovirus 1 picoBank/HPeV1/A |
| 27 | Human Parechovirus 6 |
| 28 | Human rhinovirus A89 |
| 29 | Human rhinovirus C (Strain 024) |
| 30 | Human rhinovirus B14 |
| 31 | Human enterovirus C10 strain AK11 |
| 32 | Human enterovirus C109 isolate NCA08-4327 |
| 33 | Influenza A virus (A/Zhejiang/DTIDZJU01/2013(H7N9)) |
| 34 | Influenza A virus (A/Hong Kong/1073/99(H9N2)) |
| 35 | Influenza A virus (A/Texas/5020120/99(H3N2)) |
| 36 | Influenza A virus (A/Michigan/45/2015(H1N1)) |
| 37 | Influenza B virus (B/Lee/1940) |
| 38 | Influenza B virus (B/Wisconsin/60/2008) |
| 39 | Influenza B virus (B/Brisbane 60/2008) |
| 40 | Influenza B virus (B/Colorado/ 60/2017) |
| 41 | Influenza B virus (B/Washington/02/2019) |

#### Sequence data analysis

Run metrics were evaluated on the Basespace app by analyzing cluster density and Q30 score. Sequences were then submitted for analysis to the Dragen pipeline for pathogen detection, available on the DRAGEN RNA Pathogen Detection in BaseSpace Sequence analysis (v3.5.16). In addition, sequences were also analyzed through the Dragen metagenomics pipeline (Illumina) for the detection of viruses and bacteria, in addition to the 40 viruses listed in **Table 1**.

#### Performance metric evaluation

The performance metric was calculated for both clinical and reference control samples by comparing detection results with the RT-PCR assay results. Five-performance criteria viz. positive percentage agreement (PPA), negative percentage agreement (NPA), positive predictive value (PPV), negative predictive value (NPV), accuracy, false-negative rate (FNR), and false-positive rate (FPR) were evaluated.

#### Limit of detection and reproducibility studies

The limit of detection (LoD) studies was conducted as per the FDA guidelines (https://www.fda.gov/medical-devices/coronavirus-disease-2019-covid-19-emergency-use-authorizations-medical-devices/vitro-diagnostics-euas). Briefly, SARS-CoV-2 reference material was sequentially diluted and sequenced at 10^7^ copies/ml and 10^6^ copies/ml in the singlet, 90 copies/ml, 30 copies/ml and 10 copies/ml in triplicates to evaluate both intra-run and inter-run reproducibility. The lowest concentration detected in all three triplicates was determined as the preliminary LoD. To confirm the LoD, 20 replicates of preliminary LoD were analyzed and deemed as confirmed if at least 19/20 replicates were detected.

#### Phylogenetic clustering of genomes

Inference and visualization of the phylogeny of the SARS-CoV-2 sequences were performed through the Nextstrain Command-Line Interface (CLI) tool, utilizing the associated augur and auspice toolkits [1]. To ensure proper phylogenetic inference, the following sequence exclusion criteria were applied: (i) Sequences of length less than 23,000 base pairs (∼77% of the full genome length) were excluded from the analysis, (ii) sequences without an associated metadata entry were identified and removed, and (iii) after constructing the phylogeny using a skyline coalescent method and setting a fixed clock rate of 7e-4 substitutions per site per year with a standard deviation of 2e-4, temporal outliers outside of four interquartile ranges from the root-to-tip vs time regression were removed. These steps resulted in a set of 286 sequence tips on the final tree. These parameter decisions were informed by prior analyses of SARS-CoV-2 sequences across North America [2]. These choices were validated by the resultant estimate of the Time of Most Recent Common Ancestor (TMRCA) confidence interval which spans from November 2nd, 2019 to January 26th, 2020. Similarly, the estimated clock rate of 8.55e-4 substitutions per site per year also falls within the range of estimates found throughout the literature. The metadata for the final tree has been integrated to allow for both temporal and geographical visualization in the accompanying Nextstrain instance.

## Results

### Sequencing performance

A typical sequencing run of the viral sequencing panel performed on the NextSeq550 platform consists of ∼192 samples. In this study, a total of 523 samples were sequenced in 3 runs. The first run included 143 samples sequenced using version one (V1) of the panel. Runs 2 and 3 included 380 samples sequenced using version 2 (V2) of the panel. The quality metrics and clinical performance were evaluated for the V2 of the panel that focuses less read on human control RNA genes compared to V1, resulting in a more efficient focus on pathogen RNA genes. The two runs resulted in a cluster density of 221 ± 2.1, and the Q30 scores were 90.2% and 91.3%, respectively (**Figure 1**).

**Figure 1.**
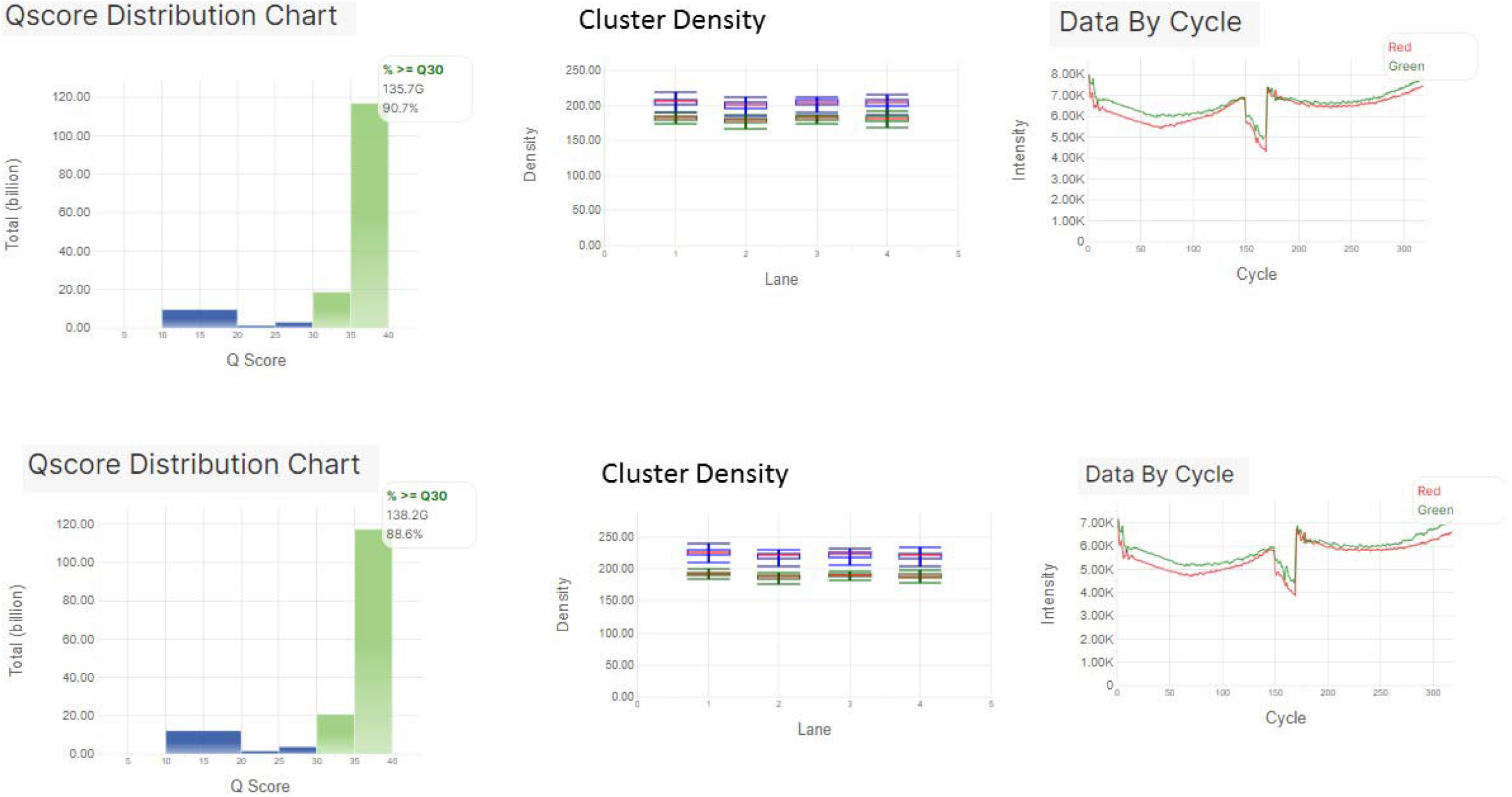
Sequencing performance of the viral sequencing panel performed on NextSeq550 platform

### Performance metric evaluation/ analytical performance

The performance metric was calculated using both the clinical samples and the reference control material. The PPA and NPA were found to be 95.98% and 85.96%, respectively. The FPR and FNR were found to be 14.04% and 4.02%, respectively. The accuracy of the assay was found to be 94.47% (**Table 2**).

**Table 2.** Performance metric evaluation of the respiratory viral panel.

| Performance Criterion | Percentage (%) |
| --- | --- |
| Positive percentage agreement (PPA) = $TP / (TP+FN)$ | 95.98 |
| Negative percentage agreement (NPA) = $TN / (TN+FP)$ | 85.96 |
| Accuracy = $TP+TN / \text{All Results}$ | 97.48 |
| False Negative Rate (FNR) = $FN / (FN+TP)$ | 4.02 |
| False Positive Rate (FPR) = $FP / (FP+TN)$ | 14.04 |

### Limit of detection and reproducibility studies

In the preliminary LoD study, all replicates were detected at the five tested concentrations using the exact biosciences reference control material. The LoD was determined to be 10 copies/ml with all 25 replicates detected. The inter-and intra-run evaluation using 90 copies/ml, 30 copies/ml and 10 copies/ml of reference material sequenced in triplicates in two different runs demonstrated high reproducibility as all replicates were detected in both runs, respectively (Figure 2). Further, all 25 replicates were detected at 10 copies/ml, demonstrating high inter-run reproducibility (Figure 3).

**Figure 2.**
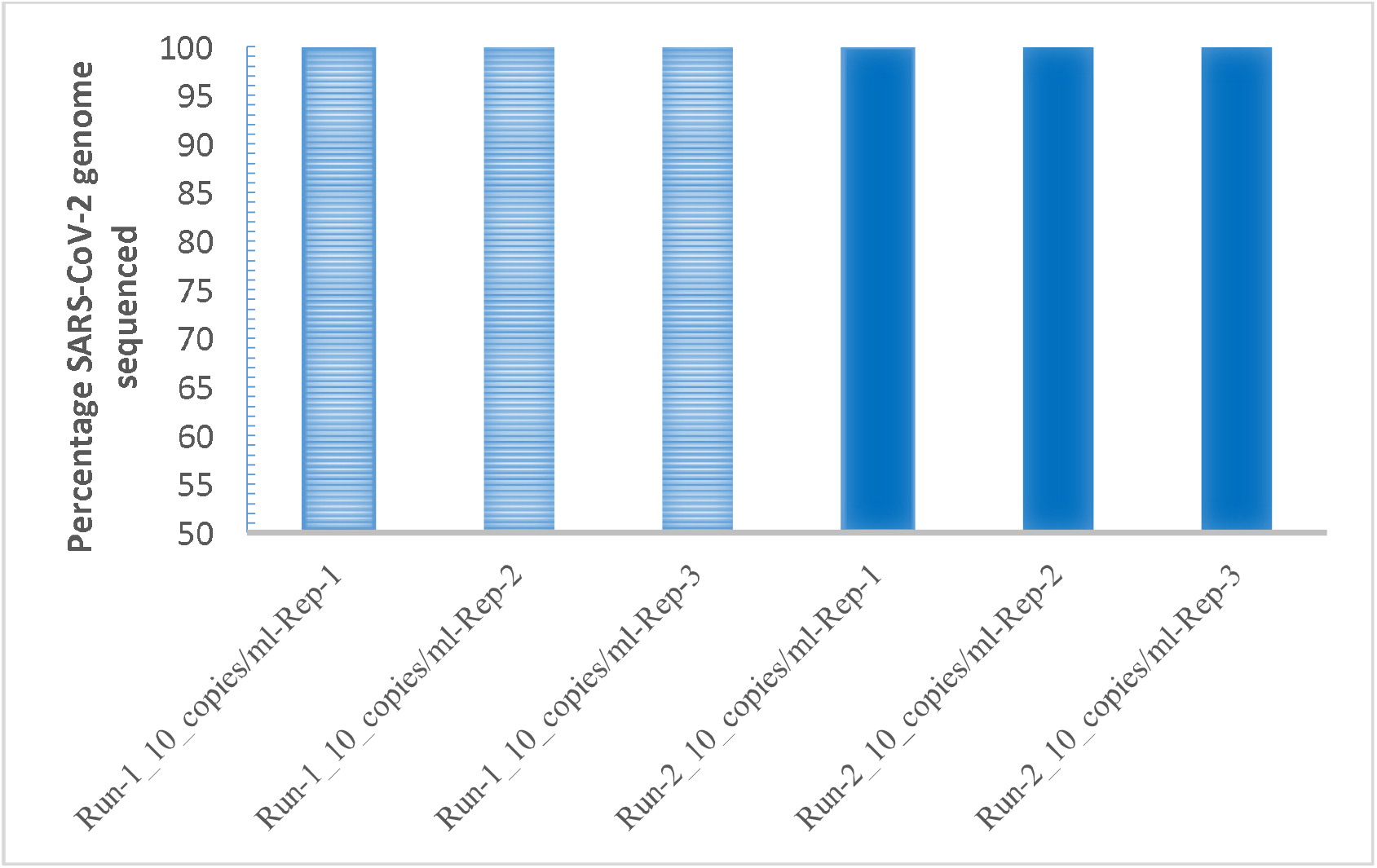
Inter-run and Intra-run performance of the RVP panel sequenced on NextSeq500/550.

**Figure 3.**
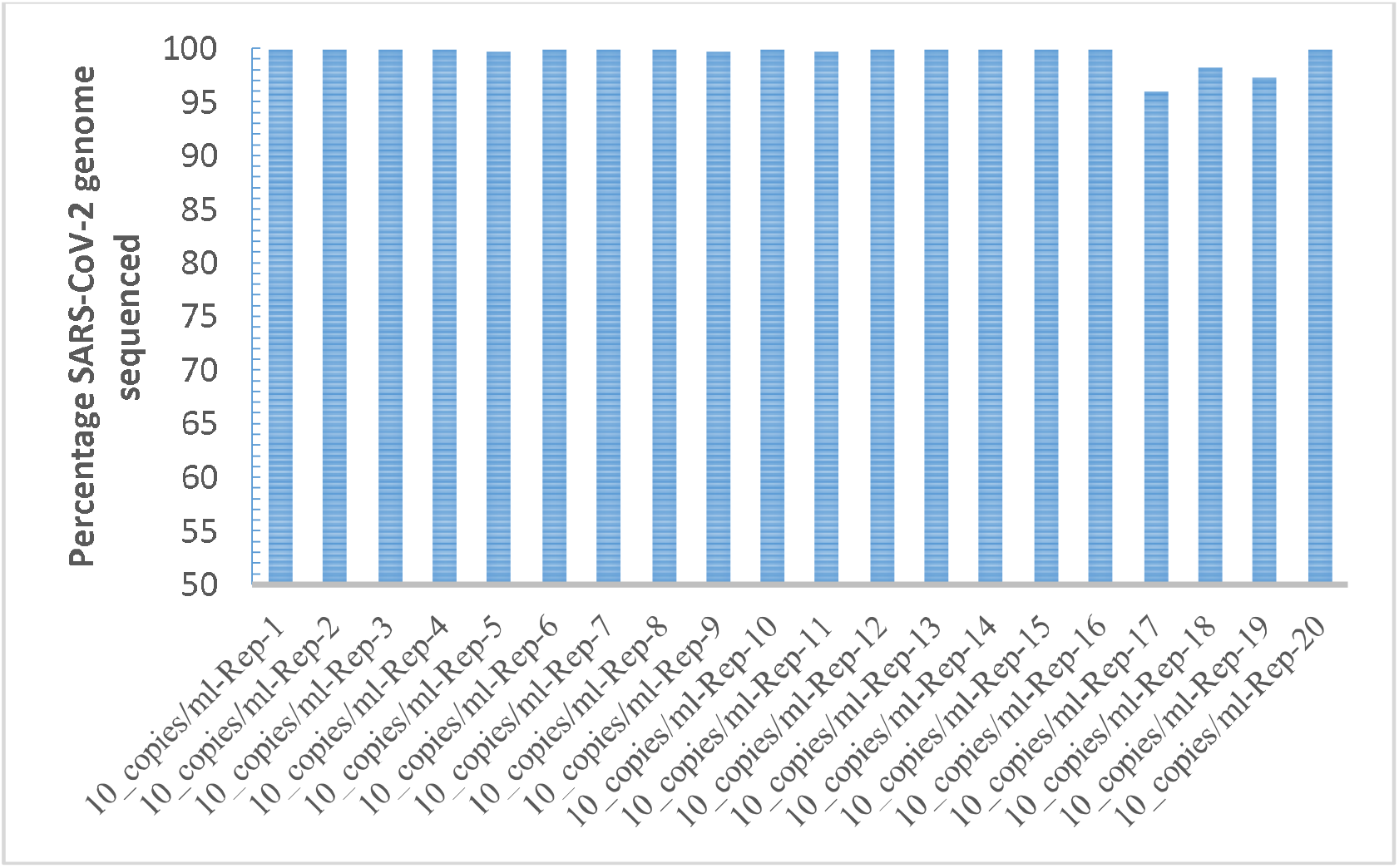
Reproducibility study using 10 copies/ml.

### Co-circulating Viruses

This study identified that 0.8% (4/483) of patients were infected with viruses other than SARS-CoV-2. Of the four patients, three had co-infection with Human enterovirus C109 isolate NICA08-4327, WU Polyomavirus, and KI polyomavirus Stockholm 60 in addition to SARS-CoV-2. One patient negative for SARS-CoV-2 was found to be infected with Human parainfluenza virus 4a.

### Phylogenetic clustering of genomes

The final instance has been posted through the Nextstrain community platform (available at https://nextstrain.org/community/Bahl-Lab-IOB/SARS_CoV2_Augusta_Edu@main). Using the open-source platform Nextstrain, we built an interactive visualization of the phylogenic analysis of our SARS-CoV-2 isolates. Two major clades for SARS-CoV-2 that were named lineage A and B in pangolin lineage can be identified from our current phylogenetic analysis. Through our Nextstrain thread, the clade for pangolin lineage B contains certain distant variants, including P4715L in ORF1ab, Q57H in ORF 3a and, S84L in ORF8 covarying with the D614G spike protein mutation were found to be the most prevalent in the early phase of the pandemic in the state of Georgia. In addition, we found the isolates from the same county form paraphyletic groups in our analysis, which indicated virus transmission between counties (**Figure 4**).

**Figure 4.**
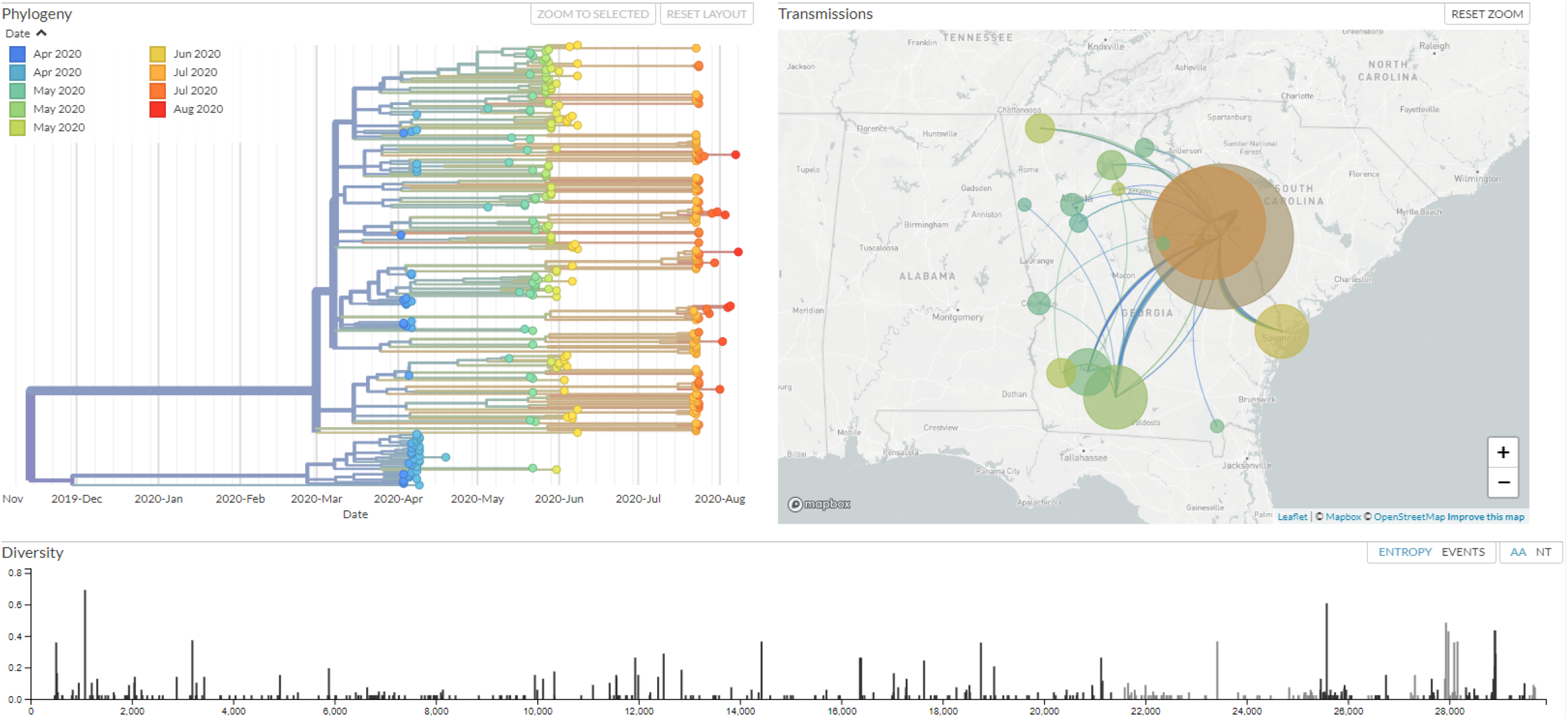
Nextstrain, interactive visualization of the phylogenic analysis of our SARS-CoV-2 isolates. (https://nextstrain.org/community/Bahl-Lab-IOB/SARS_CoV2_Augusta_Edu@main)

## Discussion

The COVID-19 pandemic has led to massive socio-economic disruption with a major impact on the health care systems across the globe. The magnitude of the pandemic required channeling almost all the available resources towards diagnosis, management, and treatment of COVID-19. The diversion of all resources to cope with the pandemic has led to two major knowledge/epidemiological/public health gaps that could further contribute to the ongoing crisis. The first is the lack of documentation of the variation in the SARS-CoV-2 genome around the world, and the second is the complete neglect of the co-circulating viruses in the population that contribute to the global disease burden. This study is an attempt to address these two major public health lacunae by evaluating the NGS-RVP, which can identify mutational variants in the SARS-CoV-2 genome and other co-circulating respiratory viruses in a single assay.

Since December of 2019, after the SARS-CoV-2 genome was characterized, various sequencing approaches have been utilized to study the virus and, for some technologies, microbiome and host responses as well. Various NGS technologies have proven their worth not just in the identification of the exact viral etiology and origin of COVID-19, but also in diagnostic assay development, vaccine design, and epidemiologic surveillance of viral transmission and evolution. By employing either shotgun metagenomics, amplicon-based or hybrid capture-based sequencing on mid-to ultra-high-throughput platforms, and single-molecule sequencing such as nanopore, complex clinical and research questions have been answered [**14-16**]. These techniques have been found to have almost comparable genome coverage, with amplicon-based approaches showing high sensitivity for low viral load samples at a lower cost. Metagenomic and hybrid capture, on the other hand, have the advantage of unbiased sequencing at a high throughput scale and co-infection identification [**17**]. In this study, we utilized an enrichment workflow, a hybrid-capture-based approach on a high throughput platform, and a metagenomic bioinformatic pipeline to interrogate patient samples that include saliva samples.

The analytical performance analysis using 522 samples demonstrated the ease of use and clinical utility of the RVP. The assay had a hands-on time of ∼10.5 hours and an assay time of ∼3 days. The sequencing run parameters met the recommended threshold values of the manufacturer, with a high consistency among samples for each parameter (both across runs and within each run). The ability to sequence both NPS and saliva samples simultaneously, adds a substantial advantage to any clinical laboratory with respect to time and efficiency.

The study evaluated 522 samples that included 483 clinical specimens and 39-reference control materials. The performance evaluation demonstrated favorable PPA, NPA, FPR, and FNR, with a high overall accuracy of the panel. The inter-and intra-run evaluation demonstrated high reproducibility, with an LoD of 10 copies/ml, rendering this assay highly sensitive and accurate for the detection of the SARS-CoV-2 genome. It is noteworthy that 88.7% (275/310) and 93.8% (291/310) of positive samples were sequenced with a coverage of more than 90% and 60% of the SARS-CoV-2 genome, respectively, that is essential for the phylogenetic clustering and mutational analysis of the genome. The coinfection rate in this study was low (0.8%) which reflects a low rate of co-infection in this population. Other studies in different populations have found variable rates of coinfection. A study in the New York metropolitan found a coinfection rate by other respiratory pathogens of less than 3% and infection by other non-SARS-CoV-2 coronaviruses of 13.1% [**18**].

Genomic sequencing and epidemiological analysis of SARS-CoV-2 are important for properly understanding the evolution and spread of this pathogen in Georgia. To this end, we build an interactive visualization of the phylogenetic analysis of our samples using the open-source platform Nextstrain, which could reflect the phylogeny and the circulating diversity during the early SARS epidemic in Georgia. Through our Nextstrain thread, we found the clade for pangolin lineage B that contains certain distant variants covarying with the D614G spike protein mutation had become increasingly prevalent as the early phase of the pandemic in the state of Georgia. Further, the isolates from the same county forming paraphyletic suggests a prolonged period of unrecognized community spreading. Such a properly maintained visualization tool can help to improve the public health efforts involving continued exploration of the evolutionary relationships between SARS-CoV-2 samples and provide a better understanding of the genetic diversity across the whole genome over time. This study demonstrates that genomic epidemiology is essential to predict disease transmission, the pattern of transmission and has the potential to recognize the imminent resurgence of a regional outbreak. Given the challenges associated with establishing such a surveillance program, it is essential to develop and maintain the infrastructure for such analysis for future pandemics.

We propose that as the COVID-19 vaccination process is underway, the screening of COVID-19 by RT-PCR-based SARS-CoV-2 detection methods needs to be complemented with two additional monitoring measures. The first is sequencing the SARS-CoV-2 genome to identify novel variants that can provide actionable information to prevent or mitigate emerging viral threats, and predict transmission/ resurgence of regional outbreaks and the second is to test for co-circulating respiratory viruses that might be independent factors contributing to the global disease burden.

## Data Availability

All relevant data is made available within the manuscript.

## Acknowledgments

The authors would like to thank Stephen Gross, Lisa Watson, Jonathan Hetzel, Eric Allen, Irina Khrebtukova, Chen Zhao, Rami Mehio, and Tim Harrington for biochemistry, analysis, and panel development work leading to the Illumina RNA Prep with Enrichment assay (cat number 20040537, Illumina) workflow with the Respiratory Virus Oligo Panel (RVOP V2) and analysis with DRAGEN RNA Pathogen Detection in BaseSpace Sequence analysis (v3.5.16).

